# Effectiveness of COVID-19 vaccines against SARS-CoV-2 variants of concern: a systematic review and meta-analysis

**DOI:** 10.1101/2021.09.23.21264048

**Authors:** Baoqi Zeng, Le Gao, Qingxin Zhou, Kai Yu, Feng Sun

## Abstract

**Background:** It was urgent and necessary to synthesize the evidence for vaccine effectiveness (VE) against SARS-CoV-2 variants of concern (VOC). We conducted a systematic review and meta-analysis to provide a comprehensive overview of the effectiveness profile of COVID-19 vaccines against VOC.

**Methods:** Published and preprinted randomized controlled trials (RCTs), cohort studies, and case-control studies that evaluated the VE against VOC (Alpha, Beta, Gamma, or Delta) were searched until 31 August 2021. Pooled estimates and 95% confidence intervals (CIs) were calculated using random-effects meta-analysis. VE was defined as (1− estimate).

**Results:** Seven RCTs (51,169 participants), 10 cohort studies (14,385,909 participants) and 16 case-control studies (734,607 cases) were included. Eight COVID-19 vaccines (mRNA-1273, BNT162b2, ChAdOx1, Ad26.COV2.S, NVX-CoV2373, BBV152, CoronaVac, and BBIBP-CorV) were included in this analysis. Full vaccination was effective against Alpha, Beta/Gamma, and Delta variants, with VE of 88.3% (95% CI, 82.4–92.2), 70.7% (95% CI, 59.9–78.5), and 71.6% (95% CI, 64.1–77.4), respectively. But partial vaccination was less effective, with VE of 59.0% (95% CI, 51.3–65.5), 49.3% (95% CI, 33.0–61.6), and 52.6% (95% CI, 43.3–60.4), respectively. mRNA vaccines seemed to have higher VE against VOC over others, significant interactions (p_interaction_ < 0.10) were observed between VE and vaccine type (mRNA vaccines vs. non-mRNA vaccines).

**Conclusions:** Full vaccination of COVID-19 vaccines is highly effective against Alpha variant, and moderate effective against Beta/Gamma and Delta variants. Partial vaccination has less VE against VOC. mRNA vaccines seem to have higher VE against Alpha, Beta/Gamma, and Delta variants over others.

## INTRODUCTION

Since emerging of coronavirus disease 2019 (COVID-19) caused by a novel severe acute respiratory syndrome coronavirus 2 (SARS-CoV-2) in December 2019, more than 225 million cases and 4.6 million deaths have been documented worldwide as of 15 September 2021 [1]. COVID-19 vaccines have been rapidly developed, and proved to be highly effective in multiple randomized clinical trials (RCTs)[2–5] and observational studies [6–8]. Most current vaccines used SARS-CoV-2 spike protein as the key antigenic target based on the originally identified Wuhan lineage virus [9]. The B.1.1.7 (Alpha) variant was first identified from genomic sequencing of samples obtained from COVID-19 patients which accounted for an expanding proportion of cases in England in late 2020 [10].Subsequently, the emergence of the B.1.351 (Beta) variant in South Africa and the P.1 (Gamma) variant in Brazil increased the COVID-19 pandemic. In December 2020, a novel SARS-CoV-2 variant, the B.1.617.2 (Delta) variant was first detected in India, causing a sharp increase in COVID-19 cases and deaths in India and surrounding countries [11].The emerging Alpha, Beta, Gamma, and Delta variants were classified as variants of concern (VOC), which were associated with the transmission increasing, more severe disease situation (e.g., increased hospitalizations or deaths), significant reduction in neutralization by antibodies generated during previous infection or vaccination, reduced effectiveness of treatments or vaccines, or diagnostic detection failures [12–17].The importance of vaccination programs and efficient public health measures will be increased if VOC have increased transmissibility or virulence.[18] It was urgent and necessary to synthesize evidence of the VE of COVID-19 vaccines against VOC. To our knowledge, there are some studies evaluating the VE of COVID-19 vaccines against VOC [19–22], but no relevant systematic review or meta-analysis has been published to date. Therefore, to gain insight in the VE of COVID-19 vaccines against VOC, we conducted a comprehensive systematic review and meta-analysis including both RCTs and observational studies. This review of the VE of COVID-19 vaccines against VOC will support global response on public health measures and vaccination programs timely and evidence based.

## METHODS

### Data sources and searches

We conducted this systematic review according to the Preferred Reporting Items for Systematic reviews and Meta-Analyses (PRISMA) guidelines [23], the protocol was registered on PROSPERO (CRD42021273986). We searched for literature published on PubMed, Embase, Cochrane Library, and the ClinicalTrials.gov website on or before 4 August 2021. Meanwhile, we also searched the medRxiv website to include eligible preprints in the last three months (from 4 May 2021 to 4 August 2021). Keywords including “COVID-19”, “SARS-CoV-2”, “vaccine”, and “variant” were used to search, the detailed search strategy was shown in the Supplementary material. Additionally, we identified references by searching the reference lists of included studies and relevant reviews. Considering that the researches on COVID-19 vaccines were updated quickly, we researched literature before submission on 31 August 2021.

### Selection of studies

We included randomized controlled trials (RCTs), cohort studies, and case-control studies that evaluated the efficacy or effectiveness of COVID-19 vaccines against VOC including B.1.1.7 (Alpha), B.1.351 (Beta), P.1 (Gamma), and B.1.617.2 (Delta). Studies enrolling general population or special populations (e.g., healthcare workers) aged 16 years or older were included. For studies that only reported VE against SARS-CoV-2 infections (without subgroup analysis of VOC), but the specific VOC accounted for 50% or more among positive tests, they were also included in the analysis. We excluded study protocols, editorials, comments, reviews, news, case reports, conference abstracts, animal studies, in vivo experiments, and analysis of antibody neutralization. Searches were limited to English articles. Preprints published in peer-reviewed journals were excluded. The primary outcome was the VE of full vaccination against VOC.

### Data extraction

Two authors reviewed titles and abstracts independently to identify eligible studies that met pre-specified inclusion criteria and extracted data. When consensus was lacking, a third reviewer was consulted. The journal name, study type, study location, vaccine information, number of participants, characteristics of subjects, and outcomes were extracted from eligible studies. We extracted SARS-CoV-2 infection information if results on both SARS-CoV-2 infection and symptomatic infection were reported. The adjusted VE or estimates of effect size (relative risks, incidence rate ratios, or odds ratios) with corresponding 95% confidence intervals (CIs) was extracted with priority. The risk of bias of RCTs was assessed using the Cochrane Collaboration’s tool [24, 25]. The risk of bias of cohort and case-control studies was assessed using the Newcastle-Ottawa scale (NOS) [26].The NOS contains 8 categories relating to methodological quality, with a maximum of 9 points. A total score of 7–9 points is considered of good quality, while a score of 4–6 points of moderate quality, and a score of 1–3 points of low quality. Two investigators reviewed the studies and judged the risk of bias.

### Statistical analysis

Pooled estimates and 95% CIs were calculated using DerSimonian and Laird random-effects meta-analysis [27]. Summary VE was defined as (1− pooled estimate) ×100%. We performed subgroup analysis stratifying by study design, vaccine type, participant, and publication. P for the difference was calculated using random-effects meta-regression, a difference between the estimates of these subgroups was considered significant if p_interaction_ < 0.10 [28]. Statistical heterogeneity between the studies was assessed with the *χ*^2^ test and the *I*^2^ statistics. *I*^2^ values of 25%, 50%, and 75% have been suggested to be indicators of low, moderate, and high heterogeneity, respectively [29]. All the analysis were performed with STATA 14.

## RESULTS

### Literature Rearch and Study characteristics

This systematic literature search identified 2226 publications and 596 preprints, after excluding duplicates and irrelevant papers, 78 published reports and 25 preprints were evaluated in full text for eligibility (Supplementary Figure 1). Finally, 33 articles (23 publications [6, 7, 19–22, 30–46] and 10 preprints [47–56]) were included in the present systematic review. There were different study designs for included studies, 7 RCTs (51,169 participants), 10 cohort studies (14,385,909 participants), and 16 case-control studies (734,607 cases). In total, 8 COVID-19 vaccines (mRNA-1273, BNT162b2, ChAdOx1, Ad26.COV2.S, NVX-CoV2373, BBV152, CoronaVac and BBIBP-CorV) and 4 VOC (Alpha, Beta, Gamma and Delta) were included in this study. BNT162b2 and mRNA-1273 are mRNA vaccines, CoronaVac, BBV152 and BBIBP-CorV are inactivated vaccines, Ad26.COV2.S and ChAdOx1 are non-replicating vector vaccines, and NVX-CoV2373 is a protein subunit vaccine. Only Ad26.COV2.S is a single-dose vaccine, therefore a one-dose regimen is regarded as full vaccination. Characteristics of individual studies are summarized in Table 1.

**Table 1.** Study characteristics and participants demographics

| First Author | Journal | Study design | VOC | Vaccine | Country | Population Characteristics | Participants |
| --- | --- | --- | --- | --- | --- | --- | --- |
| Shinde [19] | N Engl J Med | RCT (phase 2a/b) | Beta | NVX-CoV2373 | South Africa | GP; Age: 18-84 | 4,387 |
| Madhi [21] | N Engl J Med | RCT (phase 3) | Beta | ChAdOx1 | South Africa | GP; Age: 18-65 | 1,467 |
| Heath [44] | N Engl J Med | RCT (phase 3) | Alpha | NVX-CoV2373 | UK | GP; Age: 18-84 | 14,039 |
| Sadoff [45] | N Engl J Med | RCT (phase 3) | Beta | Ad26.COV2.S | South Africa | GP; Age: $\geq 18$ | 4,969 |
| Emary [22] | Lancet | RCT (phase 2/3) | Alpha | ChAdOx1 | UK | GP; Age: $\geq 18$ | 8,534 |
| Ella [48] | *Preprint* | RCT (phase 3) | Delta | BBV152 | India | GP; Age: 18-98 | 16,973 |
| Thomas [56] | *Preprint* | RCT (phase 2/3) | Beta | BNT162b2 | South Africa | GP; Age: $\geq 16$ | 800 |
| Lopez Bernal [20] | N Engl J Med | TNCC | Alpha & Delta | BNT162b2 or ChAdOx1 | UK | GP; Age: $\geq 16$ | 19,109 cases |
| Abu-Raddad [43] | N Engl J Med_ Letter | TNCC | Alpha & Beta | BNT162b2 | Qatar | GP; Age: 33 (22-40) <sup>a</sup> | 35,979 cases |
| Sheikh [39] | Lancet_ Correspondence | TNCC | Alpha & Delta | BNT162b2 or ChAdOx1 | UK | GP | 19,543 cases |
| Chemaitelly [46] | Nat Med | TNCC | Alpha & Beta | mRNA-1273 | Qatar | GP; Age: 32 (25-39) <sup>a</sup> | 66,042 cases |
| Bernal [30] | BMJ | TNCC | Alpha | BNT162b2 | UK | Older adults; Age: $\geq 70$ | 3,034 cases |
| Chung [31] | BMJ | TNCC | Alpha & Beta/Gamma | BNT162b2 and mRNA-1273 | Canada | GP; $\geq 16$ | 324,033 cases |
| Ranzani [32] | BMJ | TNCC | Gamma | CoronaVac | Brazil | Older adults; $\geq 70$ | 43,774 cases |
| Skowronski [35] | Clin Infect Dis | TNCC | Alpha & Gamma | BNT162b2 (85%) and mRNA-1273 (15%) | Canada | Older adults; $\geq 70$ | 483 cases |
| Carazo [33] | Clin Infect Dis | TNCC | Alpha | BNT162b2 and mRNA-1273 | Canada | HCWs; Age: 18-74 | 901 cases |
| Li [37] | Emerg Microbes Infect | TNCC | Delta | CoronaVac and BBIBP-CorV | China | GP; Age: 18-59 | 74 cases |
| Charmet [40] | Lancet Reg Health Eur | Case-control | Alpha & Beta/Gamma | BNT162b2 and mRNA-1273 | France | GP; Age: $\geq 20$ | 33,863 cases |
| Nasreen [51] | *Preprint* | TNCC | Alpha & Beta/Gamma | BNT162b2, mRNA-1273, or | Canada | GP; Age: $\geq 16$ | 40,828 cases |

|  |  |  | & Delta | ChAdOx1 |  |  |  |
| --- | --- | --- | --- | --- | --- | --- | --- |
| Hitchings [49] | *Preprint* | TNCC | Gamma | Corona Vac | Brazil | HCWs; Age: ≥18 | 418 cases |
| Tang [55] | *Preprint* | TNCC | Delta | BNT162b2 or mRNA-1273 | Qatar | GP; Age: 31 (24-37) <sup>a</sup> | 1,731 cases |
| Chemaitelly [47] | *Preprint* | TNCC | Alpha & Beta & Delta | BNT162b2 | Qatar | GP; Age: 32 (23-39) <sup>a</sup> | 144,075 cases |
| Pramod [53] | *Preprint* | TNCC | Delta | ChAdOx1-S | India | HCWs; Age: 34 (28-43) <sup>a</sup> | 720 cases |
| Hall [6] | Lancet | Prospective cohort | Alpha | BNT162b2 (94%) and ChAdOx1 (6%) | UK | HCWs; Age: ≥18 | 23,324 |
| Haas [38] | Lancet | Retrospective cohort | Alpha | BNT162b2 | Israel | GP; Age: ≥16 | 6,538,911 |
| Dagan [7] | N Engl J Med | Retrospective cohort | Alpha | BNT162b2 | Israel | GP; Age: ≥16 | 1,193,236 |
| Lumley [34] | Clin Infect Dis | Retrospective cohort | Alpha | BNT162b2 and ChAdOx1 | UK | HCWs; Age: 39(30-50) <sup>a</sup> | 13,109 |
| Williams [36] | Clin Infect Dis | Retrospective cohort | Gamma | mRNA-1273 | Canada | Residents and staff of LTCH | 143 |
| Nanduri [42] | Morb Mortal Wkly Rep | Retrospective cohort | Delta | mRNA-1273 or BNT162b2 | US | Nursing home residents | 5,965,607 |
| Fowlkes [41] | Morb Mortal Wkly Rep | Retrospective cohort | Delta | mRNA-1273 (33%) and BNT162b2 (65%) | US | Frontline workers | 2,840 |
| Puranik [54] | *Preprint* | Retrospective cohort | Delta (July) | mRNA-1273 or BNT162b2 | US | GP; Age: ≥18 | 67,469 |
| Lefèvre [50] | *Preprint* | Retrospective cohort | Beta | BNT162b2 | France | Residents of LTCH; Age: 89 (83-92) <sup>a</sup> | 378 |
| Pouwels [52] | *Preprint* | Retrospective cohort | Alpha & Delta | BNT162b2 or ChAdOx1 or mRNA-1273 | UK | GP; Age: ≥18 | 580,892 |

Abbreviations: VOC, variants of concern; HCWs, healthcare workers; TNCC, Test-negative case-control; LTCH, long term care homes; GP, general population; RCT, randomized controlled trial.
<sup>a</sup> Median age (interquartile range)

**Figure 1.**
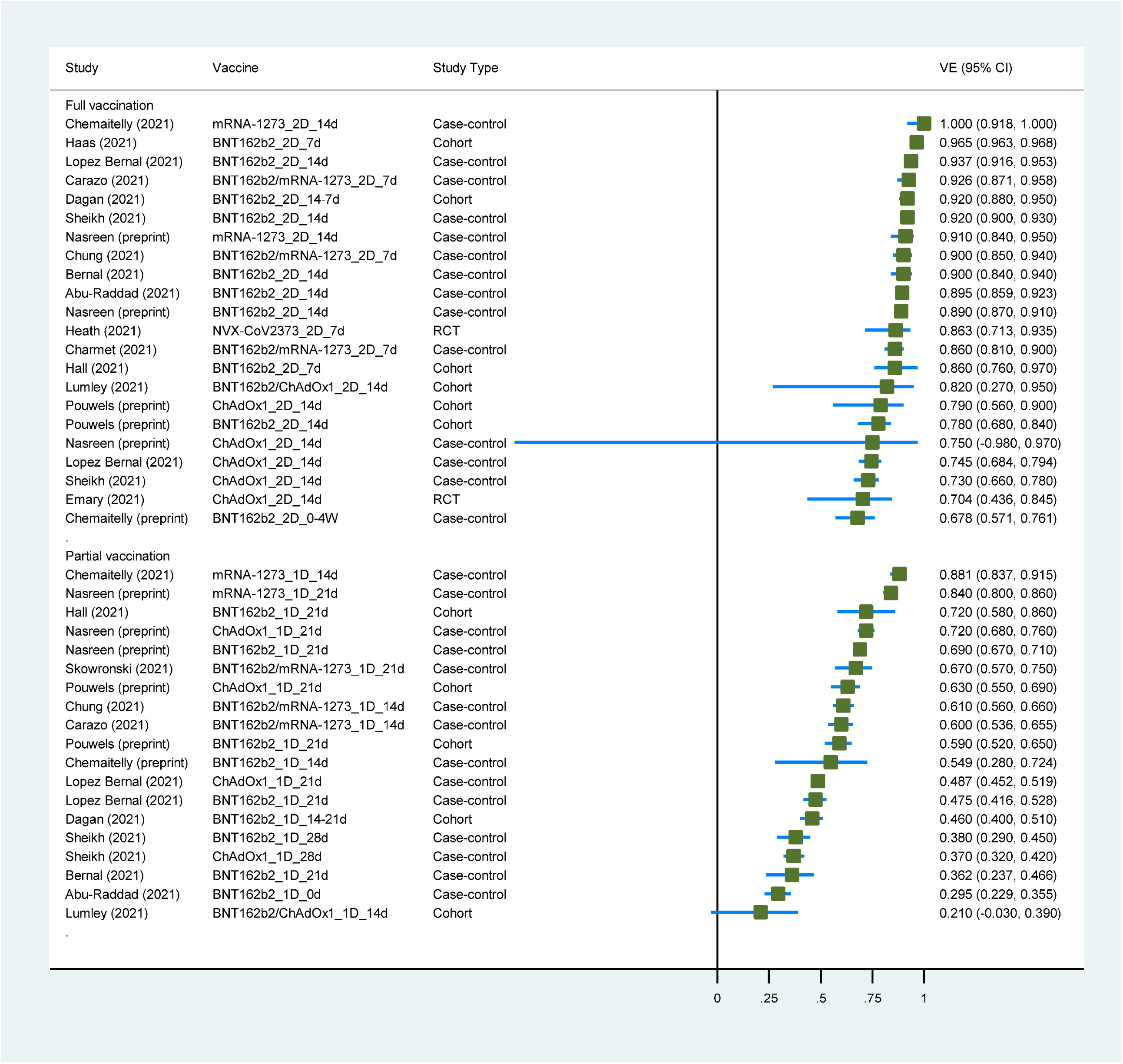
Forest plot showing vaccine effectiveness of COVID-19 vaccines against Alpah variant. Abbreviations: VE, vaccine effectiveness; CI, confidence interval; RCT, randomized controlled trial.

### Risk of Bias

All the RCTs were assessed as some concerns for overall risk-of-bias judgment. Seven of 10 cohort studies were judged as good quality, and the remaining 3 studies were moderate quality. For 16 case-control studies, 13 were considered as good quality and 3 were moderate quality. The detailed risk of bias assessment is available in online Supplementary Table 1–3.

### Vaccine Effectiveness of COVID-19 Vaccines against B.1.1.7 (Alpha) Variant

Two RCTs [22, 44], 5 cohort studies [6, 7, 34, 38, 52] and 11 case-control studies [20, 30, 31, 33, 35, 39, 40, 43, 46, 47, 51] had evaluated the VE of COVID-19 vaccines against the Alpha variant. One cohort study [52] and 2 case-control studies [47, 51] were preprints on the medRxiv website. Four COVID-19 vaccines (mRNA-1273, NVX-CoV2373, ChAdOx1 and BNT162b2) were included in this analysis. Three studies enrolled healthcare workers [6, 33, 34], two enrolled participants aged 70 or older [30, 35], and the others enrolled the general population. Characteristics of individual studies and VE for Alpha variant are summarized in Figure 1 and Supplementary Table 4.

The summary VE of full vaccination against the Alpha variant was 88.3% (95% CI, 82.4–92.2) (Table 2). Subgroup analysis by study design showed that VE was 79.4% (56.2–90.3) in 2 RCTs, 88.3% (95% CI, 70.9–95.3) in 5 cohort studies and 88.7% (95% CI, 84.2–92.0) in 10 case-control studies (p_interaction_ = 0.435). Subgroup analysis of vaccine type showed that VE was 90.9% (95% CI, 86.2–94.0) for mRNA vaccines in 15 study groups, 73.8% (95% CI, 69.8–77.4) for non-replicating vector vaccine in 5 study groups, 86.3% (95% CI, 71.3–93.5) for protein subunit vaccine in 1 study group, and 82.0% (95% CI, 27.0–95.0) for combined vaccines (BNT162b2/ChAdOx1) in 1 study group (p_interaction_ = 0.084). And we detected a significant interaction (p_interaction_ = 0.009) between VE and vaccine type (mRNA vaccines vs. non-mRNA vaccines), the VE of mRNA vaccines seemed to be higher than others. The results of subgroup analysis for participant and publication were in Table 2.

**Table 2.**
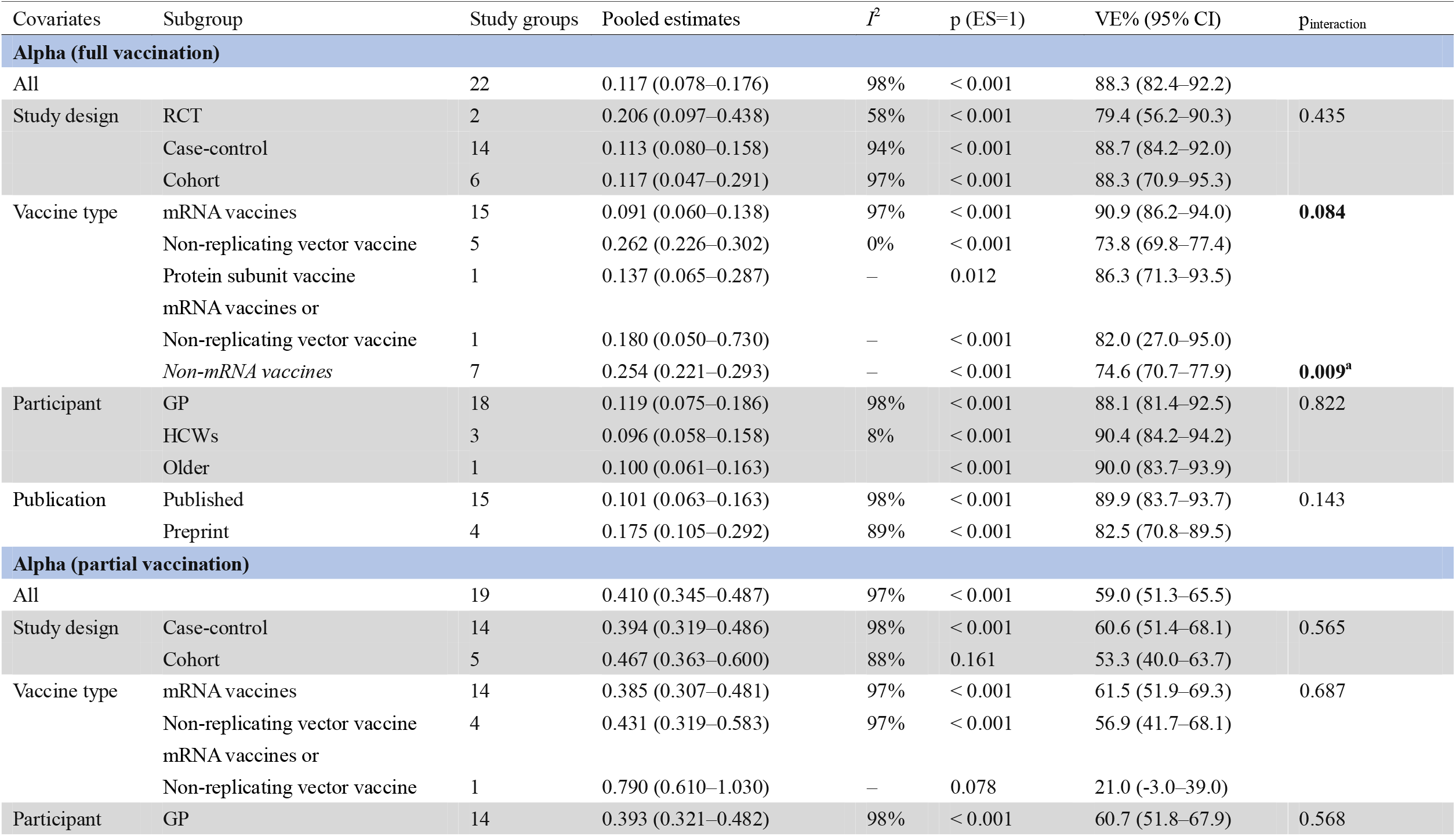
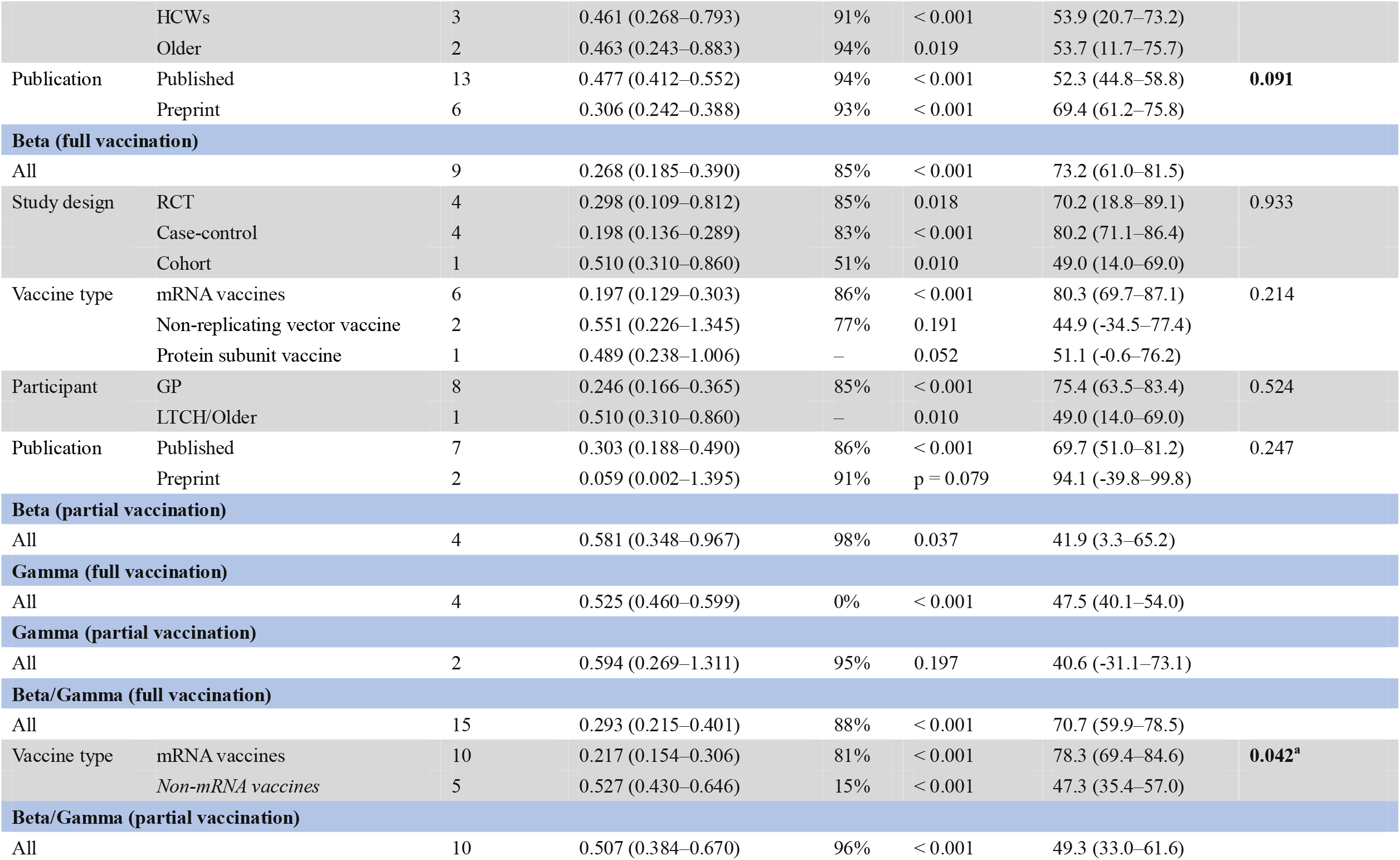
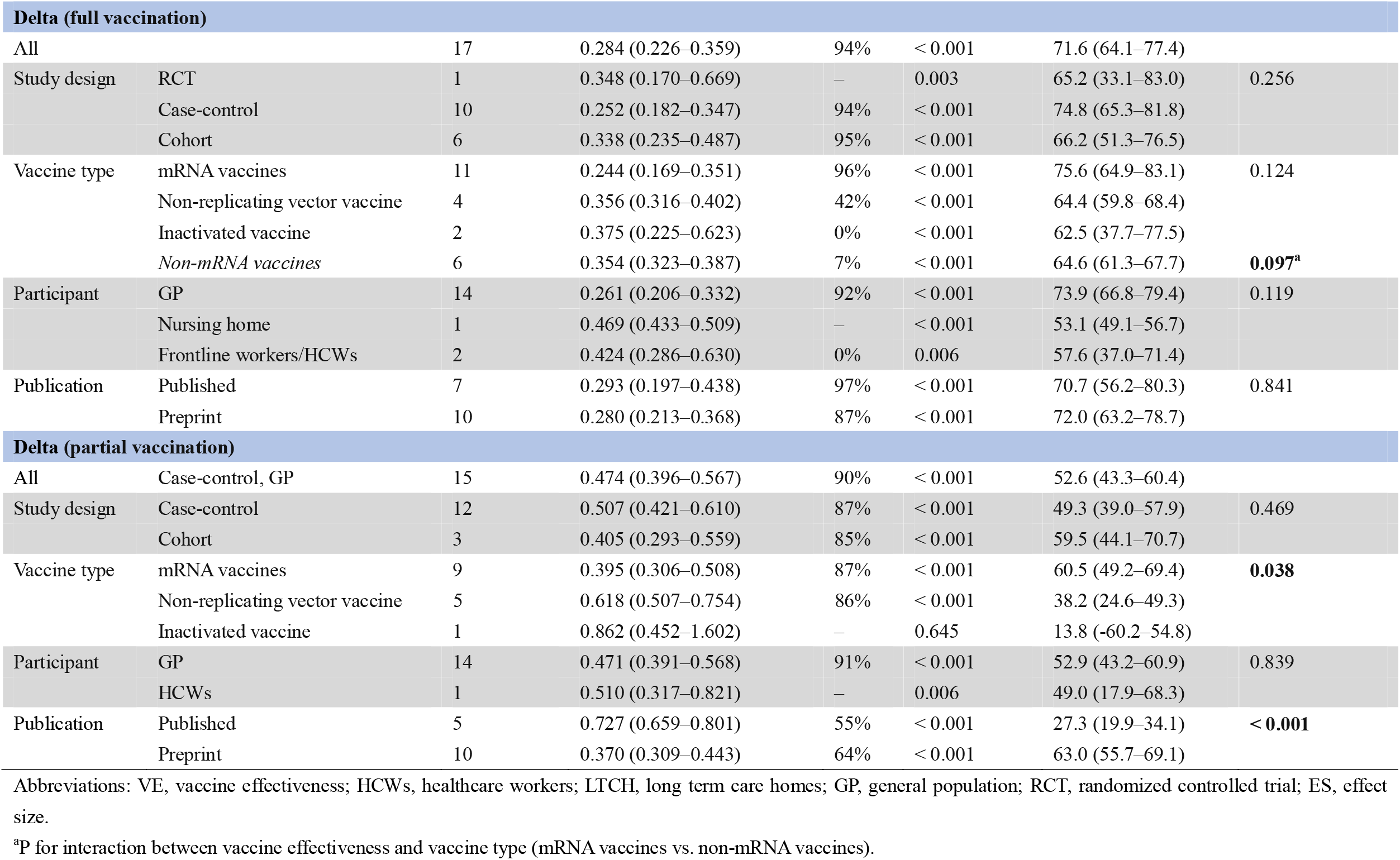
Meta-analysis and subgroup analysis for VE of COVID-19 against VOC

The summary VE of partial vaccination against the Alpha variant (Table 2) was 59.0% (95% CI, 51.3–65.5). Subgroup analysis of study design showed that VE was 53.3% (95% CI, 40.0–63.7) in 4 cohort studies and 60.6% (95% CI, 51.4–68.1) in 10 case-control studies (p_interaction_ = 0.565). Subgroup analysis by vaccine type showed that VE was 61.5% (95% CI, 51.9–69.3) for mRNA vaccines in 14 study groups, 56.9% (95% CI, 41.7–68.1) for non-replicating vector vaccine in 4 study groups, and 21.0% (95% CI, −3.0–39.0) for combined vaccines (BNT162b2/ChAdOx1) in 1 study group (p_interaction_ = 0.687).

### Vaccine Effectiveness of COVID-19 Vaccines against B.1.351 (Beta) and P.1 (Gamma) Variants

Four RCTs [21, 39, 45, 56], 1 cohort study [50], and 4 case-control studies [40, 43, 46, 47] had evaluated the VE of COVID-19 vaccines against the Beta variant. One cohort study [36] and 3 case-control studies [32, 35, 49] had evaluated the VE of COVID-19 vaccines against the Gamma variant. Both Beta and Gamma have N501Y and E484K mutations, 2 studies used a combined Beta/Gamma group because of insufficient specimens [31, 51]. One RCT [56] and 3 case-control studies [31, 49, 51] were preprints on the medRxiv website. Six COVID-19 vaccines (mRNA-1273, BNT162b2, NVX-CoV2373, ChAdOx1, CoronaVac and Ad26.COV2.S) were included in this analysis. Single-dose Ad26.COV2.S was considered a full vaccination. For the study population, 1 study enrolled health care workers [49], 2 studies enrolled participants aged 70 or older [32, 35], 2 studies enrolled participants from long-term care homes [36, 50], and the others enrolled the general population. Characteristics of individual studies and VE for Beta and Gamma variants are summarized in Figure 2 and Supplementary Table 5.

**Figure 2.**
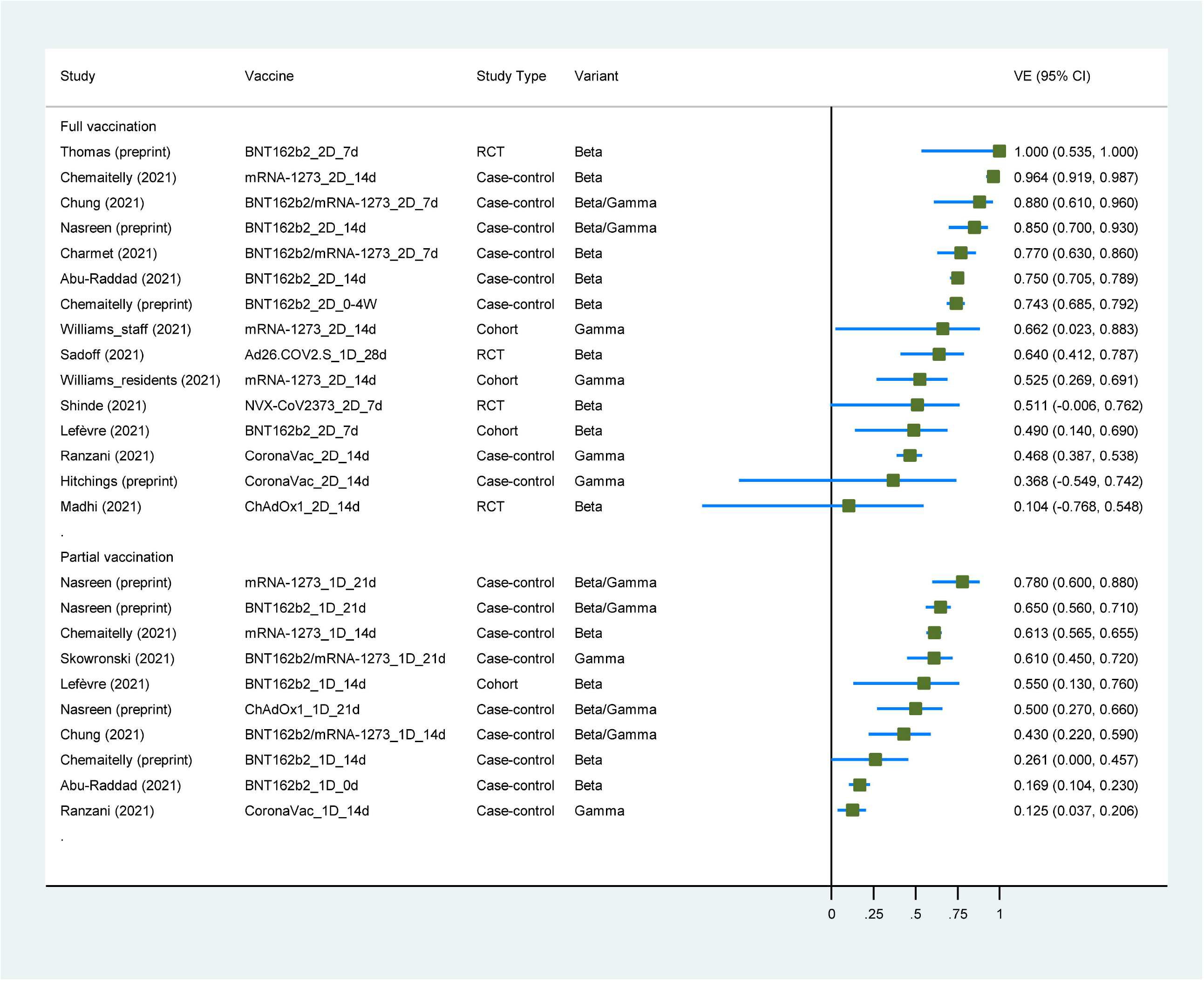
Forest plot showing vaccine effectiveness of COVID-19 vaccines against Beta/Gamma variants. Abbreviations: VE, vaccine effectiveness; CI, confidence interval; RCT, randomized controlled trial.

The summary VE of full vaccination against Beta variant were 73.2% (95% CI, 61.0–81.5) (Table 2). Subgroup analysis by study design showed that VE for Beta variant was 70.2% (95% CI, 18.8–89.1) in 4 RCTs, 80.2% (95% CI, 71.1–86.4) in 4 case-control studies, and 49.0% (95% CI, 14.0–69.0) in 1 cohort study (p_interaction_ = 0.933). Subgroup analysis of vaccine type showed that VE was 80.3% (95% CI, 69.7–87.1) for mRNA vaccines in 6 studies, 44.9% (95% CI, −34.5–77.4) for non-replicating vector vaccines in 2 studies, and 51.1% (95% CI, −0.6–76.2) for protein subunit vaccine in 1 study (p_interaction_ = 0.214). The summary VE of partial vaccination against Beta variant was 41.9% (95% CI, 3.3–65.2) in four studies.

The summary VE of full vaccination against Gamma variant was 47.5% (95% CI, 40.1–54.0) in 4 study groups, and partial vaccination against Gamma variant was 40.6% (95% CI, −31.1–73.1) in 2 study groups (Table 2). When Beta/Gamma variant was treated as one group, the summary VE of full vaccination was 70.7% (95% CI, 59.9–78.5) in 15 study groups, and partial vaccination against Gamma variant was 49.3% (95% CI, 33.0–61.6) in 10 study groups. Subgroup analysis of vaccine types showed that VE of full vaccination against Beta/Gamma was 78.3% (95% CI, 69.4–84.6) for mRNA vaccines in 10 study groups and 47.3% (95% CI, 35.4–57.0) for non-mRNA vaccines in 5 study groups (p_interaction_ = 0.042), the VE for mRNA vaccines seemed to be higher than others. The results are in Table 2.

### Vaccine Effectiveness of COVID-19 Vaccines against B.1.617.2 (Delta) Variant

One RCT [48], 4 cohort studies [41, 42, 52, 54] and 7 case-control studies [20, 37, 39, 47, 51, 53, 55] had evaluated the VE of COVID-19 vaccines against the Delta variant. Seven studies are preprints on the medRxiv website [47, 48, 51–55]. Six COVID-19 vaccines (mRNA-1273, BBV152, ChAdOx1, BNT162b2, CoronaVac and BBIBP-CorV) were included in this analysis. Two studies enrolled health care workers or frontline workers [41, 53], 1 study enrolled participants in the nursing home [42], and the others enrolled the general population. Characteristics of individual studies and VE for Delta variant are summarized in Figure 3 and Supplementary Table 6.

**Figure 3.**
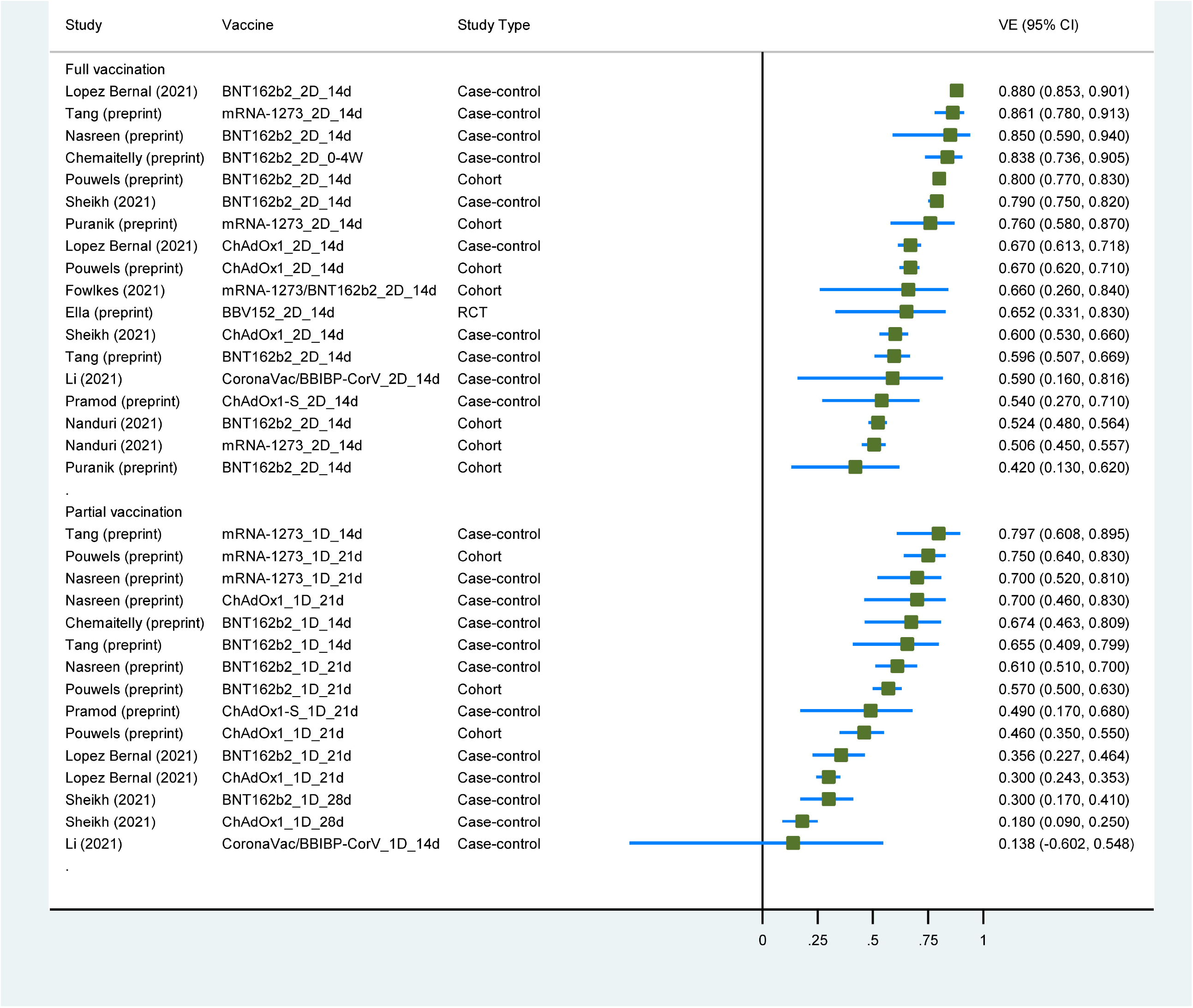
Forest plot showing vaccine effectiveness of COVID-19 vaccines against Delta variant. Abbreviations: VE, vaccine effectiveness; CI, confidence interval; RCT, randomized controlled trial.

The summary VE of full vaccination against the Delta variant was 71.6% (95% CI, 64.1–77.4) (Table 2). Subgroup analysis of study design showed that VE for Delta variant was 65.2% (95% CI, 33.1–83.0) in 1 RCT, 74.8% (95% CI, 65.3–81.8) in 7 case-control studies, and 66.2% (95% CI, 51.3–76.5) in 4 cohort studies (p_interaction_ = 0.256). Subgroup analysis by vaccine type showed that VE was 75.6% (95% CI, 64.9–83.1) for mRNA vaccines in 11 study groups, 64.4% (95% CI, 59.8–68.4) for non-replicating vector vaccine in 4 study groups, and 62.5% (95% CI, 37.7–77.5) for inactivated vaccines in 2 study groups (p_interaction_ = 0.124). An interaction (p_interaction_ = 0.097) between VE and vaccine type (mRNA vaccines vs. non-mRNA vaccines) was found, the VE for mRNA vaccines seemed to be higher than others.

The summary VE of partial vaccination against the Delta variant was 52.6% (95% CI, 43.3–60.4) (Table 2). Subgroup analysis for study design showed that VE for Delta variant was 59.5% (95% CI, 44.1–70.7) in 1 cohort study, and 49.3% (95% CI, 39.0–57.9) in 7 case-control studies (p_interaction_ = 0.469). Subgroup analysis of vaccine type showed that VE was 60.5% (95% CI, 49.2–69.4) for mRNA vaccines in 9 study groups, 38.2% (95% CI, 24.6–49.3) for non-replicating vector vaccine in 5 study groups, and 13.8% (95% CI, 60.2–54.8) for inactivated vaccines in 1 study group (p_interaction_ = 0.038). The VE for mRNA vaccines seemed to be higher than others.

## DISCUSSION

We conducted this systematic review and meta-analysis to synthesize evidence on the VE of COVID-19 vaccines against VOC during the pandemic. This study has four main findings. First, full vaccination of COVID-19 vaccines was effective against Alpha, Beta/Gamma and Delta variants, with the VE of 88.3% (95% CI, 82.4–92.2), 70.7% (95% CI, 59.9–78.5) and 71.6% (95% CI, 64.1–77.4), respectively. Second, partial vaccination has lower VE against Alpha, Beta/Gamma and Delta variants, with the VE of 59.0% (95% CI, 51.3–65.5), 49.3% (95% CI, 33.0–61.6), 52.6% (95% CI, 43.3–60.4), respectively. Third, mRNA vaccines (BNT162b2 or mRNA-1273) seemed to have higher VE against VOC over other vaccines. Fourth, more evidence was needed to evaluate the VE of COVID-19 against the Gamma and Delta variants. To our knowledge, our study is the first comprehensive systematic review and meta-analysis to characterize the VE of COVID-19 vaccines against VOC.

The evidence for the Gamma variant was insufficient, only four studies were included [32, 35, 36, 49]. One study [36] enrolled residents or staff in long term care homes and two [32, 35] enrolled older adults aged 70 years and older, which reflected the VE against Gamma variant for elderly and frail people. The remaining one study evaluating the CoronaVac vaccine against the Gamma variant enrolled healthcare workers, but unmeasured confounding which led to downward bias in the VE estimate was reported [49].

The main results in this study were in consistent with a recent meta-analysis for neutralizing antibodies against SARS-CoV-2 variants, which showed that Alpha, Beta, Gamma and Delta variants significantly escaped natural-infection-mediated neutralization, with an average of 1.4-fold, 4.1-fold, 1.8-fold, and 3.2-fold reduction in live virus neutralization assay [57]. Despite the reduction in neutralization titers against Alpha variant, they remain robust, and there is no evidence of vaccine escape in one study [58]. Escape of Beta variant from neutralization by convalescent plasma and vaccine-induced sera was observed in some studies [12, 59, 60]. Although neutralization titers against Gamma variant are reduced, it is hoped that immunization with vaccines designed against parent strains will protect Gamma variant infection [61]. The Delta variant escapes neutralization by some antibodies that target the receptor-binding domain or N-terminal domain, the neutralization titers against Delta was three to five folds than Alpha variant when two-dose of the vaccine administrated.[14] This study also supports the two-dose vaccine regimen recommended by the FDA and EMA, which is consistent with an in-vitro study for SARS-CoV-2 variants of concern [62].

This review included 3 study designs evaluating 8 COVID-19 vaccines against 4 VOC in different populations. There is high heterogeneity between studies, and high statistical heterogeneity is also observed in most analysis. Other factors like the definition of outcomes (all SARS-CoV-2 or symptomatic infection), days after vaccination and participant’s characteristics (e.g., age and race) may also contribute to the heterogeneity. Therefore, we mainly performed narrative descriptive synthesis.

This systematic review and meta-analysis has some limitations. First, 21% of studies (7 of 33) are nonrandomized. The imbalance between groups in observational studies is a concern, so potential selection bias may be existent. Second, 30% of studies (10 of 33) are preprints and have not been certified by peer review. We may not identify errors in data analysis or reporting due to the lack of a rigorous vetting process. Third, although we performed qualitative analysis by different stratifications, heterogeneity was still high in most quantitative analysis. Forth, VE against hospitalization or death related to VOC is not included in our analysis. Finally, the evidence of COVID-19 vaccines against Gamma and Delta variants is not enough, more research is needed in the future.

## CONCLUSION

Full vaccination of COVID-19 vaccines is highly effective against Alpha variant, and moderate effective against Beta/Gamma and Delta variant. Partial vaccination has less effectiveness against all VOC. Therefore, full vaccination is recommended against variants of SARS-CoV-2. mRNA vaccines (BNT162b2 or mRNA-1273) seem to have higher VE against Alpha, Beta/Gamma, or Delta over other vaccines, but more evidence is needed to confirm this finding.

## Data Availability

The data for the article is available

## Notes

### Author contributions

KY and FS conceived the study. BZ and KY designed the study. BZ, LG and QZ undertook the literature review and extracted the data. BZ and QZ coded the statistical analysis, figures, and appendix. BZ and LG interpreted the data and wrote the first draft of the manuscript. All authors reviewed and revised subsequent drafts and approved the final version.

