## Supplementary Material for "Effectiveness of COVID-19 vaccines against SARS-CoV-2 variants of concern: a systematic review and meta-analysis"

### Search strategy (August 31, 2021)

**Search strategy for Embase (934 items)**

1. 'coronavirus disease 2019'/exp OR 'Severe acute respiratory syndrome coronavirus 2'/exp OR (COVID-19 OR SARS-CoV-2):ti,ab
2. vaccine/exp OR vaccination/exp OR immunization/exp OR (vaccin* OR immuniz*):ti,ab
3. (variant* OR B.1.1.7 OR B.1.351 OR P.1 OR B.1.617.2):ti,ab
4. #1 AND #2 AND #3

**Search strategy for Pubmed (1203 items)**

1. COVID-19[Mesh] OR SARS-CoV-2[Mesh] OR "COVID-19 Vaccines"[Mesh] OR COVID-19[Title/Abstract] OR SARS-CoV-2[Title/Abstract]
2. vaccines[Mesh] OR vaccination[Mesh] OR immunization[Mesh] OR vaccin*[Title/Abstract] OR immuniz*[Title/Abstract]
3. variant*[Title/Abstract] OR B.1.1.7[Title/Abstract] OR B.1.351[Title/Abstract] OR P.1[Title/Abstract] OR B.1.617.2[Title/Abstract]
4. #1 OR #2 OR #3

**Search strategy for Cochrane Library (23 items)**

1. [mh COVID-19]
2. [mh SARS-CoV-2]
3. COVID-19:ti,ab
4. SARS-CoV-2:ti,ab
5. #1 OR #2 OR #3 OR #4
6. [mh vaccines]
7. [mh vaccination]
8. [mh immunization]
9. vaccin*:ti,ab
10. immuniz*:ti,ab
11. #6 OR #7 OR #8 OR #9 OR #10
12. (variant* OR B.1.1.7 OR B.1.351 OR P.1 OR B.1.617.2):ti,ab
13. #5 AND #11 AND #12

**Search strategy for Clinicaltrial.gov (66 items)**

(vaccines OR vaccine OR vaccination OR immunization) AND (variant OR variants) AND (COVID-19 OR SARS-CoV-2)

**Search strategy for medRxiv (596 items)**

(vaccin*) AND (variant*) AND (COVID-19 OR SARS-CoV-2) AND (Efficacy OR Effectiveness)

### Supplementary Figure

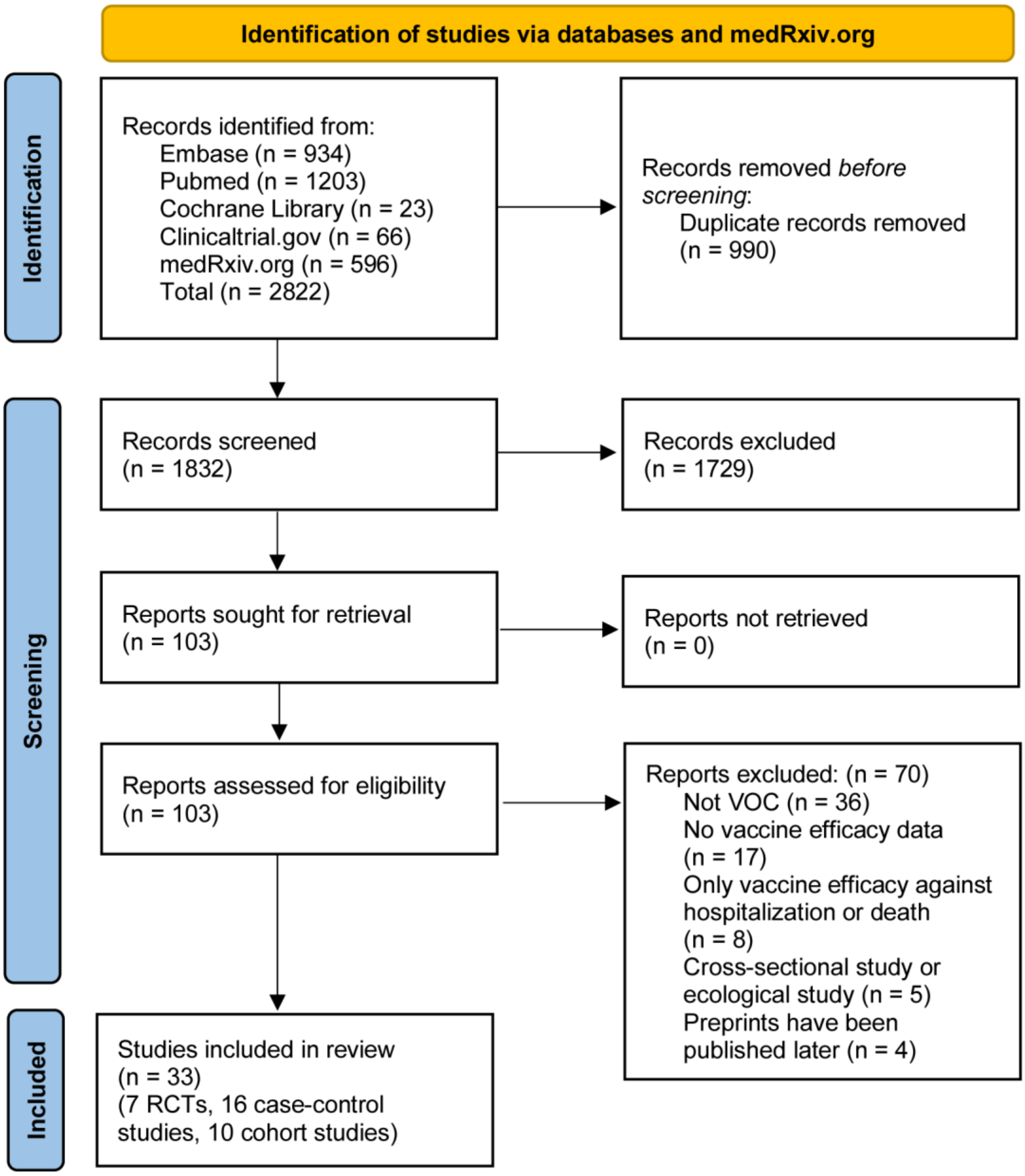


**Supplementary Figure 1.** Flow chart of literature search and study selection.

### Supplementary Tables

**Supplementary Table 1**. Risk of bias for included RCTs

| Study (First author) | Randomization process | Deviations from intended interventions | Missing outcome data | Measurement of the outcome | Selection of the reported result | **Overall bias** |
|---|---|---|---|---|---|---|
| Heath | Low | Some concerns | Some concerns | Low | Some concerns | Some concerns |
| Emary | Low | Some concerns | Some concerns | Low | Some concerns | Some concerns |
| Shinde | Low | Low | Some concerns | Low | Some concerns | Some concerns |
| Madhi | Low | Some concerns | Low | Low | Some concerns | Some concerns |
| Sadoff | Low | Some concerns | Some concerns | Low | Low | Some concerns |
| Thomas (preprint) | Low | Some concerns | Low | Low | Low | Some concerns |
| Ella (preprint) | Low | Some concerns | Some concerns | Low | Low | Some concerns |

**Supplementary Table 2**. Risk of bias for included cohort studies

| | Selection | | | | Comparability | Exposure | | | | |
|---|---|---|---|---|---|---|---|---|---|---|
| | Item 1 | Item 2 | Item 3 | Item 4 | Item 5 | Item 6 | Item 7 | Item 7 | Total score | **Quality** |
| Hall | 1 | 1 | 1 | 1 | 1 | 1 | 1 | 1 | 8 | Good |
| Haas | 1 | 1 | 1 | 0 | 1 | 1 | 1 | 1 | 7 | Good |
| Lumley | 1 | 1 | 1 | 0 | 1 | 1 | 1 | 1 | 7 | Good |
| Williams | 0 | 1 | 1 | 0 | 0 | 0 | 1 | 1 | 4 | Moderate |
| Nanduri | 1 | 1 | 1 | 0 | 1 | 1 | 1 | 1 | 7 | Good |
| Fowlkes | 0 | 1 | 1 | 0 | 1 | 0 | 1 | 1 | 5 | Moderate |
| Puranik (preprint) | 1 | 1 | 1 | 0 | 2 | 1 | 1 | 1 | 8 | Good |
| Lefèvre (preprint) | 0 | 1 | 1 | 0 | 1 | 1 | 1 | 1 | 6 | Moderate |
| Dagan | 1 | 1 | 1 | 0 | 2 | 1 | 1 | 1 | 8 | Good |
| Pouwels (preprint) | 1 | 1 | 1 | 0 | 2 | 1 | 1 | 1 | 8 | Good |

Item 1: Representativeness of the exposed cohort.

Item 2: Selection of the non-exposed cohort.

Item 3: Ascertainment of exposure.

Item 4: Demonstration that outcome of interest was not present at start of study.

Item 5: Comparability of cohorts on the basis of the design or analysis.

Item 6: Assessment of outcome.

Item 7: Was follow up long enough for outcomes to occur.

Item 8: Adequacy of follow up of cohorts.

**Supplementary Table 3**. Risk of bias for included case-control studies

| | Selection | | | | Comparability | Exposure | | | | |
|---|---|---|---|---|---|---|---|---|---|---|
| | Item 1 | Item 2 | Item 3 | Item 4 | Item 5 | Item 6 | Item 7 | Item 7 | Total score | **Quality** |
| Charmet | 1 | 1 | 1 | 1 | 1 | 1 | 0 | 1 | 7 | Good |
| Nasreen (preprint) | 1 | 1 | 1 | 1 | 1 | 1 | 1 | 1 | 8 | Good |
| Skowronski | 1 | 0 | 1 | 1 | 1 | 1 | 1 | 1 | 7 | Good |
| Hitchings (preprint) | 1 | 0 | 1 | 1 | 0 | 1 | 1 | 1 | 6 | Moderate |
| Ranzani | 1 | 0 | 1 | 1 | 2 | 1 | 1 | 1 | 8 | Good |
| Lopez Bernal | 1 | 1 | 1 | 1 | 2 | 1 | 1 | 1 | 9 | Good |
| Tang (preprint) | 1 | 1 | 1 | 1 | 2 | 1 | 1 | 1 | 9 | Good |
| Li | 1 | 0 | 1 | 1 | 1 | 1 | 1 | 1 | 7 | Good |
| Abu-Raddad | 1 | 1 | 1 | 1 | 2 | 1 | 1 | 1 | 9 | Good |
| Sheikh | 1 | 0 | 1 | 1 | 0 | 1 | 1 | 1 | 6 | Moderate |
| Chemaitelly | 1 | 1 | 1 | 1 | 2 | 1 | 1 | 1 | 9 | Good |
| Bernal | 1 | 0 | 1 | 1 | 1 | 1 | 1 | 1 | 7 | Good |
| Chung | 1 | 1 | 1 | 1 | 1 | 1 | 1 | 1 | 8 | Good |
| Carazo | 1 | 0 | 1 | 1 | 1 | 1 | 1 | 1 | 7 | Good |
| Chemaitelly (preprint) | 1 | 1 | 1 | 1 | 2 | 1 | 1 | 1 | 9 | Good |
| Pramod (preprint) | 1 | 0 | 1 | 1 | 1 | 1 | 1 | 0 | 6 | Moderate |

Item 1: Is the case definition adequate?

Item 2: Representativeness of the cases.

Item 3: Selection of controls.

Item 4: Definition of controls.

Item 5: Comparability of cases and controls on the basis of the design or analysis.

Item 6: Ascertainment of exposure.

Item 7: Same method of ascertainment for cases and controls.

Item 8: Non-Response Rate.

**Supplementary Table 4.** VE of COVID-19 vaccines against B.1.1.7 (Alpha) variant

| First author/Country | Study design | Vaccine/control or Cases/controls | Participants | Age (years) | Vaccine | Outcomes | Day_1 | VE% of one dose (95% CI) | Day_2 | VE% of two doses (95% CI) | Variants |
|---|---|---|---|---|---|---|---|---|---|---|---|
| Heath/UK | RCT | 7,020/7,020 | GP | 18-84 | NVX-CoV2373 | Symptomatic | NA | NA | 7 | 86.3 (71.3–93.5) | Alpha |
| Emary/UK | RCT | 4,244/4,290 | GP | ≥18 | ChAdOx1 | Symptomatic | NA | NA | 14 | 70.4 (43.6–84.5) | Alpha |
| Hall/UK | Cohort | 8,203/15,121 | HCWs | ≥18 | BNT162b2 | All infections | 21 | 72.0 (58.0–86.0) | 7 | 86.0 (76.0–97.0) | Alpha (>50%) |
| Haas/Israel | Cohort | 4,714,932/1,823,979 | GP | ≥16 | BNT162b2 | All infections | NA | NA | 7 | 96.5 (96.3–96.8) | Alpha (95%) |
| Lumley/UK | Cohort | 10,651/10,513 | HCWs | 39 (30-50) [1] | BNT162b2/ ChAdOx1 | All infections | 14 | 21.0 (-3.0–39.0) | 14 | 82.0 (27.0–95.0) | Alpha (51%) |
| Dagan/Israel | Cohort | 596,618/596,618 | GP | ≥16 | BNT162b2 | All infections | 14-21 | 46.0 (40.0–51.0) | 7 | 92.0 (88.0–95.0) | Alpha (80%) |
| Pouwels/UK _*preprint* | Cohort | 384,543 (total) | GP | ≥18 | BNT162b2 | All infections | 21 | 59.0 (52.0–65.0) | 14 | 78.0 (68.0–84.0) | Alpha (dominant) |
| | | | GP | ≥18 | ChAdOx1 | All infections | 21 | 63.0 (55.0–69.0) | 14 | 79.0 (56.0–90.0) | Alpha |
| Lopez Bernal/UK | TNCC | 9,682/171,834 | GP | ≥16 | BNT162b2/ ChAdOx1 | Symptomatic | 21 | 48.7 (45.5–51.7) | 14 | 87.5 (85.1–89.5) | Alpha |
| | | 7,812/120,761 | GP | ≥16 | BNT162b2 | Symptomatic | 21 | 47.5 (41.6–52.8) | 14 | 93.7 (91.6–95.3) | Alpha |
| | | 9,183/147,444 | GP | ≥16 | ChAdOx1 | Symptomatic | 21 | 48.7 (45.2–51.9) | 14 | 74.5 (68.4–79.4) | Alpha |
| Abu-Raddad/Qatar | TNCC | 19,017/19,432 | GP | 33 (22-40) [1] | BNT162b2 | All infections | 0 | 29.5 (22.9–35.5) | 14 | 89.5 (85.9–92.3) | Alpha |
| Bernal/UK | TNCC | 3,034/9,487 | Older | ≥70 | BNT162b2 | Symptomatic | 21 | 36.2 (23.7–46.6) | 14 | 90.0 (84.0–94.0) | Alpha |
| Chemaitelly/Qatar | TNCC | 23,904/23,951 | GP | 32 (25-39) [1] | mRNA-1273 | All infections | 14 | 88.1 (83.7–91.5) | 14 | 100 (91.8–100) | Alpha |
| Charmet/France | Case-cont rol | 31,313/3,644 | GP | ≥20 | BNT162b2/ mRNA-1273 | All infections | NA | NA | 7 | 86.0 (81.0–90.0) | Alpha |
| Skowronski/Canada | TNCC | 303/10,388 | Older | ≥70 | BNT162b2/ mRNA-1273 | All infections | 21 | 67.0 (57.0–75.0) | NA | NA | Alpha |
| Sheikh/UK | TNCC | 6,205/187,318 | GP | NA | BNT162b2 | All infections | 28 | 38.0 (29.0–45.0) | 14 | 92.0 (90.0–93.0) | Alpha |
| | | 6,884/203,579 | GP | NA | ChAdOx1 | All infections | 28 | 37.0 (32.0–42.0) | 14 | 73.0 (66.0–78.0) | Alpha |
| Carazo/Canada | TNCC | 901/46,862 | HCWs | 18-74 | BNT162b2/ mRNA-1273 | All infections | 14 | 60.0 (53.6–65.5) | 7 | 92.6 (87.1–95.8) | Alpha |

| | | | | | | | | | | | |
|---|---|---|---|---|---|---|---|---|---|---|---|
| Chung/Canada | TNCC | 12,582/259,986 | GP | ≥16 | BNT162b2/ mRNA-1273 | Symptomatic | 14 | 61.0 (56.0–66.0) | 7 | 90.0 (85.0–94.0) | Alpha |
| Chemaitelly/Qatar *_preprint* | TNCC | 1,888/1,888 | GP | 32 (23-39) [1] | BNT162b2 | All infections | 14 | 54.9 (28.0–72.4) | 0-4 weeks | 67.8 (57.1–76.1) | Alpha |
| Nasreen/Canada *_preprint* | TNCC | 368,323/51,540 | GP | ≥16 | BNT162b2 | Symptomatic | 21 | 69.0 (67.0–71.0) | 14 | 89.0 (87.0–91.0) | Alpha |
| | | | GP | ≥16 | mRNA-1273 | Symptomatic | 21 | 84.0 (80.0–86.0) | 14 | 91.0 (84.0–95.0) | Alpha |
| | | | GP | ≥16 | ChAdOx1 | Symptomatic | 21 | 72.0 (68.0–76.0) | 14 | 75.0 (-98.0–97.0) | Alpha |

Abbreviations: VE, vaccine effectiveness; HCWs, healthcare workers; TNCC, test-negative case-control; GP, general population; Day_1, days after the 1$^{st}$ dose; Day_2, days after the 2$^{nd}$ dose; RCT, randomized controlled trial; CI, confidence interval.

[1] Median age (interquartile range)

### Supplementary Table 5. VE of COVID-19 vaccines against B.1.351 (Beta) and P.1 (Gamma) variant

| First author/Country | Study design | Vaccine/control or Cases/controls | Participants | Age (years) | Vaccine | Outcomes | Day_1 | VE% of one dose (95% CI) | Day_2 | VE% of two doses (95% CI) | Variants |
|---|---|---|---|---|---|---|---|---|---|---|---|
| Shinde/South Africa | RCT | 1,357/1,327 | GP | 18-84 | NVX-CoV2373 | Symptomatic | NA | NA | NA | 51.1 (-0.6–76.2) | Beta |
| Madhi/South Africa | RCT | 750/714 | GP | 18-65 | ChAdOx1 | Symptomatic | NA | NA | NA | 10.4 (-76.8–54.8) | Beta |
| Sadoff/South Africa | RCT | 2,473/2,496 | GP | ≥18 | Ad26.COV2.S | Symptomatic | 14 | 52.0 (30.3–67.4) | NA | NA | Beta |
| | RCT | 2,473/2,496 | GP | ≥18 | Ad26.COV2.S | Symptomatic | 28 | 64.0 (41.2–78.7) | NA | NA | Beta |
| Thomas/South Africa_*preprint* | RCT | 291/276 | GP | ≥16 | BNT162b2 | All infections | NA | NA | 7 | 100 (53.5–100) | Beta |
| Abu-Raddad/Qatar | TNCC | 21,685/22,204 | GP | 33 (22-40)[1] | BNT162b2 | All infections | 0 | 16.9 (10.4–23.0) | 14 | 75.0 (70.5–78.9) | Beta |
| Chemaitelly/Qatar | TNCC | 48,297/48,456 | GP | 32 (25-39)[1] | mRNA-1273 | All infections | 14 | 61.3 (56.5–65.5) | 14 | 96.4 (91.9–98.7) | Beta |
| Charmet/France | Case-control | 2,550/3,644 | GP | ≥20 | BNT162b2/ mRNA-1273 | All infections | NA | NA | 7 | 77.0 (63.0–86.0) | Beta (95.1%) Gamma (4.9%) |
| Lefèvre/France | Cohort | 338/40 | LTCH | 89 (83-92)[1] | BNT162b2 | All infections | 14 | 55.0 (13.0–76.0) | 7 | 49.0 (14.0–69.0) | Beta |
| Chemaitelly/Qatar_*preprint* | TNCC | 3,277/3,277 | GP | 32 (23-39)[1] | BNT162b2 | All infections | 14 | 26.1 (0.0-45.7) | 0-4 weeks | 74.3 (68.5-79.2) | Beta |
| Williams/Canada | Cohort | 48/12 | Residents of LTCH | NA | mRNA-1273 | All infections | NA | NA | 14 | 52.5 (26.9–69.1) | Gamma |
| | | 43/40 | Staff of LTCH | NA | mRNA-1273 | All infections | NA | NA | 14 | 66.2 (2.3–88.3) | Gamma |
| Ranzani/Brazil | TNCC | 26,433/17,622 | Older | ≥70 | CoronaVac | Symptomatic | 14 | 12.5 (3.7–20.6) | 14 | 46.8 (38.7–53.8) | Gamma |
| Skowronski/Canada | TNCC | 180/10,388 | Older | ≥70 | BNT162b2/ mRNA-1273 | All infections | 21 | 61.0 (45.0–72.0) | NA | NA | Gamma |
| Hitchings/Brazil_*preprint* | TNCC | 418/418 | HCWs | ≥18 | CoronaVac | Symptomatic | NA | NA | 14 | 36.8 (-54.9–74.2) | Gamma (66%) |
| Chung/ Canada | TNCC | NA/259,986 | GP | ≥16 | BNT162b2/ mRNA-1273 | Symptomatic | 14 | 43.0 (22.0–59.0) | 7 | 88.0 (61.0–96.0) | Beta/Gamma |

| | | | | | | | | | | | |
|---|---|---|---|---|---|---|---|---|---|---|---|
| Nasreen/Canada_*preprint* | TNCC | 3,005/351,540 | GP | ≥16 | BNT162b2 | Symptomatic | 21 | 65.0 (56.0–71.0) | 14 | 85.0 (70.0–93.0) | Beta/Gamma |
| | | | GP | ≥16 | mRNA-1273 | Symptomatic | 21 | 78.0 (60.0–88.0) | NA | NA | Beta/Gamma |
| | | | GP | ≥16 | ChAdOx1 | Symptomatic | 21 | 50.0 (27.0–66.0) | NA | NA | Beta/Gamma |

Abbreviations: VE, vaccine effectiveness; HCWs, healthcare workers; TNCC, test-negative case-control; LTCH, long term care homes; GP, general population; Day_1, days after the 1st dose; Day_2, days after the 2nd dose; RCT, randomized controlled trial; CI, confidence interval.

[1] Median age (interquartile range)

### Supplementary Table 6. VE of COVID-19 vaccines against B.1.617.2 (Delta) variant

| First author/ Country | Study design | Vaccine/control or Cases/controls | Participants | Age (years) | Vaccine | Outcomes | Day_1 | VE% of one dose (95% CI) | Day_2 | VE% of two doses (95% CI) | Variants |
|---|---|---|---|---|---|---|---|---|---|---|---|
| Ella/India/_preprint | RCT | 8,471/8,502 | GP | 18-98 | BBV152 | Symptomatic | NA | NA | 14 | 65.2 (33.1–83.0) | Delta |
| Lopez Bernal/UK | TNCC | 5,876/171,834 | GP | ≥16 | BNT162b2/ ChAdOx1 | Symptomatic | 21 | 30.7 (25.2–35.7) | 14 | 79.6 (76.7–82.1) | Delta |
| | | 4,302/120,761 | GP | ≥16 | BNT162b2 | Symptomatic | 21 | 35.6 (22.7–46.4) | 14 | 88.0 (85.3–90.1) | Delta |
| | | 5,617/147,444 | GP | ≥16 | ChAdOx1 | Symptomatic | 21 | 30.0 (24.3–35.3) | 14 | 67.0 (61.3–71.8) | Delta |
| Li/China | TNCC | 74/292 | GP | 18-59 | CoronaVac/ CNBG | All infections | 14 | 13.8 (-60.2–54.8) | 14 | 59.0 (16.0–81.6) | Delta |
| Nanduri/US | Cohort | 5,011,746/953,861 | Nursing home | NA | BNT162b2/ mRNA-1273 | All infections | NA | NA | 14 | 53.1 (49.1–56.7) | Delta |
| | | 3,248,732/953,861 | | NA | BNT162b2 | All infections | NA | NA | 14 | 52.4 (48.0–56.4) | Delta |
| | | 1,763,014/953,861 | | NA | mRNA-1273 | All infections | NA | NA | 14 | 50.6 (45.0–55.7) | Delta |
| Fowlkes/US | Cohort | 2,352/488 | Frontline workers | NA | mRNA-1273/ BNT162b2 | All infections | NA | NA | 14 | 66.0 (26.0–84.0) | Delta |
| Sheikh/UK | TNCC | 4,043/185,156 | GP | NA | BNT162b2 | All infections | 28 | 30.0 (17.0–41.0) | 14 | 79.0 (75.0–82.0) | Delta |
| | | 4,679/201,374 | GP | NA | ChAdOx1 | All infections | 28 | 18.0 (9.0–25.0) | 14 | 60.0 (53.0–66.0) | Delta |
| Puranik/US_*preprint* | Cohort | 21,079/24,444 | GP | ≥18 | mRNA-1273 | All infections | NA | NA | 14 | 76.0 (58.0–87.0) | Delta (70%) |
| | | 21,946/24,444 | GP | ≥18 | BNT162b2 | All infections | NA | NA | 14 | 42.0 (13.0–62.0) | Delta (70%) |
| Chemaitelly/Qatar _*preprint* | TNCC | 1,935/1,935 | GP | 32 (23-39) | BNT162b2 | All infections | 14 | 67.4 (46.3–80.9) | 0-4 weeks | 83.8 (73.6–90.5) | Delta |
| Nasreen/Canada _*preprint* | TNCC | 991/351,540 | GP | ≥16 | BNT162b2 | Symptomatic | 21 | 61.0 (51.0–70.0) | 14 | 85.0 (59.0–94.0) | Delta |
| | | | GP | ≥16 | mRNA-1273 | Symptomatic | 21 | 70.0 (52.0–81.0) | NA | NA | Delta |
| | | | GP | ≥16 | ChAdOx1 | Symptomatic | 21 | 70.0 (46.0–83.0) | NA | NA | Delta |
| Tang/Qatar | TNCC | 1,959/1,959 | GP | 31 (24-37) | BNT162b2/ | All infections | 14 | 70.5 (55.2–80.6) | 14 | 63.3 (55.7–69.6) | Delta |

| | | | | | | | | | | | |
|---|---|---|---|---|---|---|---|---|---|---|---|
| *_preprint* | | | | | mRNA-1273 | | | | | | |
| | | 1,830/1,830 | GP | 31 (24-37) | BNT162b2 | All infections | 14 | 65.5 (40.9–79.9) | 14 | 59.6 (50.7–66.9) | Delta |
| | | 1,666/1,666 | GP | 31 (24-37) | mRNA-1273 | All infections | 14 | 79.7 (60.8–89.5) | 14 | 86.1 (78.0–91.3) | Delta |
| Pramod/India | TNCC | 360/360 | HCWs | 34 (28-43) | ChAdOx1-S | All infections | 21 | 49.0 (17.0–68.0) | 14 | 54.0 (27.0–71.0) | Delta |
| *_preprint* | | | | | | | | | | | |
| Pouwels/UK | Cohort | 358,983 (total) | GP | ≥18 | BNT162b2 | All infections | 21 | 57.0 (50.0–63.0) | 14 | 80.0 (77.0–83.0) | Delta |
| *_preprint* | | | | | | | | | | | (dominant) |
| | | | GP | ≥18 | ChAdOx1 | All infections | 21 | 46.0 (35.0–55.0) | 14 | 67.0 (62.0–71.0) | Delta |
| | | | GP | 18–64 | mRNA-1273 | All infections | 21 | 75.0 (64.0–83.0) | NA | NA | Delta |

Abbreviations: VE, vaccine effectiveness; HCWs, healthcare workers; TNCC, test-negative case-control; LTCH, long term care homes; GP, general population; Day_1, days after the 1st dose; Day_2, days after the 2nd dose; RCT, randomized controlled trial; CI, confidence interval.

[1] Median age (interquartile range)
